# Risk Factors and Long-term Prognosis for Coinfection of Nontuberculous Mycobacterial Pulmonary Disease and Chronic Pulmonary Aspergillosis: A Multicenter Observational Study in Japan

**DOI:** 10.1101/2025.02.03.25320241

**Authors:** Yasuhiro Tanaka, Shotaro Ide, Takahiro Takazono, Kazuaki Takeda, Naoki Iwanaga, Masataka Yoshida, Naoki Hosogaya, Yusei Tsukamoto, Satoshi Irifune, Takayuki Suyama, Tomo Mihara, Akira Kondo, Tsutomu Kobayashi, Yuichi Fukuda, Eisuke Sasaki, Toyomitsu Sawai, Yasuhito Higashiyama, Kohji Hashiguchi, Minako Hanaka, Toshihiko Ii, Kiyoyasu Fukushima, Kosaku Komiya, Taiga Miyazaki, Kazuhiro Yatera, Koichi Izumikawa, Akitsugu Furumoto, Katsunori Yanagihara, Hiroshi Mukae

## Abstract

**Background and objective:** Nontuberculous mycobacterial pulmonary disease (NTM-PD) is a chronic respiratory infection with increasing prevalence and mortality worldwide. Chronic pulmonary aspergillosis (CPA) is a significant complication of NTM-PD and is associated with a poor prognosis. In this multicenter, retrospective, cohort study, we examined the epidemiology, comorbidities, risk factors for CPA coinfection, and long-term prognoses of patients with NTM-PD infected with CPA in Japan.

**Methods:** Patients aged ≥ 8 years with newly diagnosed NTM-PD who visited 18 acute-care hospitals in Kyushu, Japan, between 2010 and 2017 were included. Medical records were reviewed for patient characteristics, underlying diseases, mycobacterial species, laboratory data, radiological features, *Aspergillus* coinfection, and all-cause mortality. Risk factors for CPA coinfection were analyzed using multiple logistic regression, and survival analysis was performed before and after propensity score matching with risk factors.

**Results:** Among 1,304 patients with NTM-PD, 45 (3.5%) were diagnosed with CPA during the observation period. Risk factors for CPA coinfection included male sex, chronic obstructive pulmonary disease, oral corticosteroid use, and cavity formation. All-cause mortality was significantly higher in patients with NTM-PD with CPA than in those without CPA (log-rank test, *P* < .001; crude HR, 3.98). Survival analysis after propensity score matching confirmed that CPA was an independent poor prognostic factor (log-rank test, *P* = .036; adjusted HR, 1.59).

**Conclusion:** CPA is an independent poor prognostic factor in patients with NTM-PD. Clinicians must consider CPA when treating patients with NTM-PD, particularly those with high-risk factors, to ensure their timely diagnosis and management.

**Summary at a Glance:** A study of 1,304 NTM-PD patients in Japan found that 3.5% had chronic pulmonary aspergillosis (CPA), significantly increasing their mortality risk. Key risk factors for CPA included male sex, COPD, corticosteroid use, and cavity formation. CPA is an independent poor prognostic factor, highlighting the need for timely diagnosis and management.

## INTRODUCTION

Nontuberculous mycobacteria (NTM) are ubiquitous environmental microorganisms that cause various infections in humans. Nontuberculous mycobacterial pulmonary disease (NTM-PD) is a chronic respiratory infection caused by NTM that has been recognized as an important respiratory infection in recent years. Recent reports have indicated a worldwide increase in NTM-PD prevalence, with Japan showing particularly high rates and increasing mortality.^1–4^ *Mycobacterium avium* and *Mycobacterium intracellulare* are the most common NTM species, accounting for 90% of NTM-PD cases in Japan.^2,5^ NTM-PD requires a long-term multidrug treatment regimen; however, it is sometimes refractory and recurs after treatment.^6^

Recently, chronic pulmonary aspergillosis (CPA) has been identified as an important complication of NTM-PD, which typically leads to a poor prognosis.^7–9^ While both NTM-PD and CPA individually require long-term treatment, coinfection makes treatment difficult because of the drug-drug interactions between rifamycin, macrolides, and azole antifungal agents.^10^ Moreover, CPA is associated with the exacerbation of chronic infections and increased mortality rates in patients with NTM-PD. Previous studies have elucidated the risk factors, comorbidity rates, and prognostic implications of CPA in patients with NTM-PD.^7–9,11^ However, a substantial portion of this knowledge stems from investigations conducted at single centers or studies with constrained sample sizes. Therefore, we conducted a multicenter, retrospective, cohort study of patients with NTM-PD in Kyushu, Japan, including both tertiary care and community hospitals. We aimed to determine the epidemiology, comorbidities, risk factors, and long-term prognoses of patients with NTM-PD who were coinfected with CPA.

## METHODS

### Study Design

This multicenter, retrospective, observational, cohort study was conducted at 18 acute care hospitals in Kyushu, Japan (Table S1): Nagasaki, Fukuoka, Oita, Miyazaki, and Saga (NFOMS NTM study). Patients aged ≥18 years with newly diagnosed NTM-PD who visited the study centers between January 1, 2010, and December 31, 2017, were included. The exclusion criteria were death during the collection of specimens for diagnosis and confirmation of diagnosis, referral to another institution for initial treatment after diagnosis, and patients whose last observation was before the date of confirmed diagnosis. Medical records were reviewed by respiratory medicine specialists at each facility for patient characteristics, underlying diseases, isolated mycobacterial species, laboratory data, and radiological features when the patient was definitively diagnosed with NTM-PD, chronic infections, treatments, and outcomes during the observation period. The patients were followed up until December 31, 2022. All data were collected using Research Electronic Data Capture (REDCap^®^). This study was conducted in accordance with the guidelines of the Declaration of Helsinki and approved by the Institutional Review Board of Nagasaki University Hospital (approval number: 22121903). Verbal informed consent was obtained whenever possible, and as this was a retrospective observational study, an opt-out procedure was provided to patients.

### Definitions for NTM-PD

The Japanese Society for Tuberculosis and Nontuberculous Mycobacteriosis (JSTNM) 2008 criteria were used to diagnose NTM-PD.^12^ The diagnostic criteria for JSTNM are congruent with those of the American Thoracic Society/European Respiratory Society/European Society of Clinical Microbiology and Infectious Diseases/Infectious Diseases Society of America Clinical Practice Guideline 2020. However, the JSTNM criteria do not include clinical symptoms.^6,13^ NTM species identification methods vary according to the institution and include PCR for *M. avium* and *M. intracellulare*, DNA-DNA hybridization, or matrix-assisted laser desorption/ionization-time-of-flight mass spectrometry. Radiological classification was based on chest computed tomography (CT) and classified by experts as a nodular-bronchiectatic (NB) pattern, NB with cavity pattern, fibrocavitary pattern, single nodule, hypersensitivity pneumonitis, and others.

### Diagnosis for CPA

CPA was diagnosed based on the criteria of the Japanese Domestic Guidelines for Management of Deep-seated Mycosis 2014.^14^ Briefly, it included clinical symptoms; radiological findings; serological tests, including *Aspergillus* antibody, *Aspergillus* galactomannan antigen, and β-1,3-D-glucan; and proof of *Aspergillus* species using pathology or culture. These diagnostic criteria included CPA such as simple aspergilloma, chronic cavitary pulmonary aspergillosis (CCPA), chronic fibrosing pulmonary aspergillosis (CFPA), *Aspergillus* nodule, and subacute invasive pulmonary aspergillosis (SAIA) according to the European Respiratory Society/European Society of Clinical Microbiology and Infectious Diseases guidelines.^15^ As per Japanese guidelines, CCPA, CFPA, and SAIA were comprehensively included as chronic progressive pulmonary aspergillosis. Patients with *Aspergillus* spp. colonization were excluded from CPA.

### Statistical Analysis

Risk factors for NTM-PD with CPA were analyzed using the Mann–Whitney U test for continuous variables and Fisher’s exact test for nominal variables. Multiple logistic regression analysis was performed by selecting items deemed clinically important by the experts. Risk factors with a frequency of <5 were excluded from the analysis. Propensity score matching was performed between patients with NTM-PD without CPA and those with NTM-PD with CPA, using the results obtained from multiple logistic regression analysis. Nearest-neighbor matching at a ratio of 1:5 was performed using a caliper of 0.2. A standardized difference of <0.10 indicated sufficient balance between the groups. Survival analysis was performed for the NTM-PD with and without CPA groups using the log-rank test and Cox proportional hazards model before and after propensity score matching. All *P*-values were two-sided, and *P*-values ≤ 5 were considered statistically significant. All statistical analyses were performed using EZR (version 1.67; Saitama Medical Center, Jichi Medical University, Saitama, Japan), which is a graphical user interface for R (version 4.3.3; The R Foundation for Statistical Computing, Vienna, Austria).^16^ It is a modified version of the R commander designed to add statistical functions frequently used in biostatistics.

## RESULTS

### Patient Characteristics

During the study period, 1,317 patients were newly diagnosed with NTM-PD, and 1,304 appropriate cases were evaluated (Figure 1). The median observation period was 59 months (interquartile range (IQR), 19–95 months). Patient characteristics are shown in Table 1. Of these, 45 patients (3.5%) were diagnosed with CPA during the observation period: 12 patients were diagnosed with CPA before NTM-PD diagnosis, 13 were diagnosed with CPA concurrently with NTM-PD, and 20 were diagnosed with CPA after NTM-PD diagnosis. In the CPA subtype, there were 3 cases of simple aspergilloma and 42 cases of chronic progressive pulmonary aspergillosis, including CCPA, CFPA, and SAIA.

**FIGURE 1.**
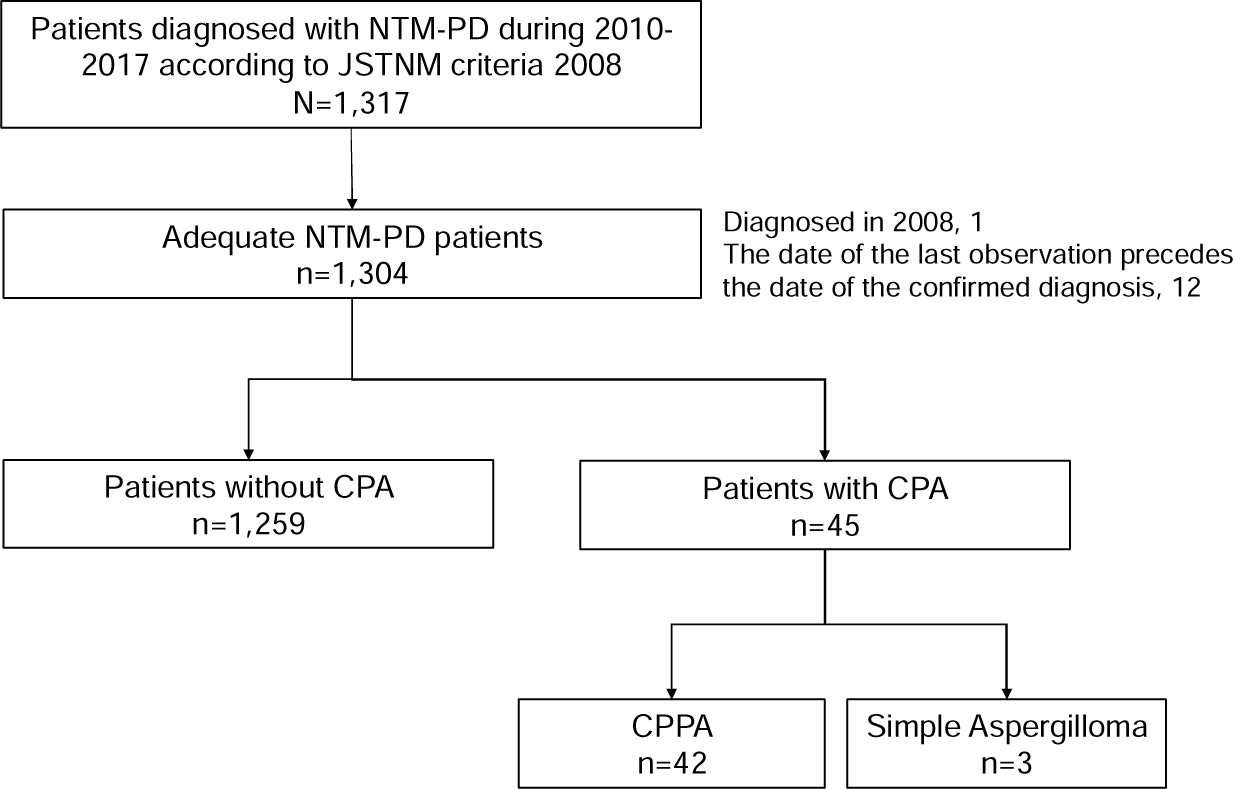
Flowchart of study patients. During the study period, 1,317 patients were newly diagnosed with NTM-PD, and 1,304 appropriate cases were evaluated. The median observation period was 59 months (IQR, 19–95 months). Of these, 45 patients (3.5%) were diagnosed with CPA, of which there were 3 cases of simple aspergilloma and 42 cases of CPPA, including CCPA, CFPA, and SAIA. CCPA, chronic cavitary pulmonary aspergillosis; CFPA, chronic fibrosing pulmonary aspergillosis; CPA, chronic pulmonary aspergillosis; CPPA, chronic progressive pulmonary aspergillosis; JSTNM, The Japanese Society for Tuberculosis and Nontuberculous Mycobacteriosis; NTM-PD, nontuberculous mycobacterial pulmonary disease; SAIA, subacute invasive pulmonary aspergillosis.

**TABLE 1.** Characteristics of Patients With Nontuberculous Mycobacterial Pulmonary Disease at Diagnosis.

|  | N =<br>1,304 (%) |
| --- | --- |
| Age (y), mean (SD) | 70.7 ± 11.5 |
| Sex, male | 423 (32.4) |
| Smoking habit | 310 (23.8) |
| Body mass index (kg/cm<sup>2</sup>) (n = 1,090), mean (SD) | 19.6±3.1 |
| Pulmonary comorbidities |  |
| Old tuberculosis | 353 (27.1) |
| Bronchiectasis | 100 (7.7) |
| Interstitial lung disease | 99 (7.6) |
| Lung tumor | 67 (5.1) |
| COPD | 49 (3.8) |
| Pulmonary emphysema | 45 (3.5) |
| Bronchial asthma | 43 (3.3) |
| Chronic bronchitis | 38 (2.9) |
| Post lung operation | 33 (2.5) |
| Pneumoconiosis | 25 (1.9) |
| Pulmonary aspergillosis | 16 (1.2) |
| Chronic respiratory failure with home oxygen therapy | 13 (1.0) |
| Asbestos-related pleural disease | 4 (0.3) |
| Pulmonary sarcoidosis | 4 (0.3) |
| Systemic comorbidities |  |
| Solid tumor, localized | 160 (12.3) |
| Connective tissue diseases | 113 (8.7) |
| Diabetes mellitus | 97 (7.4) |
| Cerebrovascular disease | 47 (3.6) |
| Liver disease, moderate/severe | 43 (3.3) |
| Congestive heart failure | 33 (2.5) |
| Solid tumor, metastatic | 33 (2.5) |
| Myocardial infarction | 31 (2.4) |
| Dementia | 29 (2.2) |
| Lymphoma | 24 (1.8) |
| Peptic ulcer disease | 19 (1.5) |
| Renal disease | 18 (1.4) |
| Peripheral vascular disease | 7 (0.5) |
| Leukemia | 5 (0.4) |
| AIDS | 3 (0.2) |
| Paralysis | 2 (0.2) |
| Medications for comorbidities | 166 (12.7) |
| Oral corticosteroids | 98 (7.5) |
| Inhaled corticosteroids | 67 (5.1) |
| Immunosuppressants | 27 (2.1) |
| Chemotherapy | 18 (1.4) |
| Biological agents | 8 (0.6) |
| Laboratory Data, mean (SD) |  |
| Albumin (g/dL) (n = 1,111) | 3.8 ± 0.7 |
| ESR (mm/h) (n = 532) | 34 ± 26 |
| White blood cell count (/μL) (n = 1,226) | 6,275 ± 2773 |
| Lymphocyte count (/μL) (n = 1,215) | 1,429 ± 704 |
| Galactomannan antigen (COI) (n = 303) | 0.72 ± 0.93 |
| Radiological features (n = 1,200) |  |
| Bronchiectasis | 990 (75.9) |
| Cavity formation | 343 (26.3) |
| Cavity formation ≥ 2 cm | 192 (14.7) |
| Cavity formation ≥ 2 lobes | 112 (8.6) |
| Radiological patterns (n = 1,200) |  |
| Non-cavitary NB pattern | 785 (60.2) |
| Cavitary NB pattern | 241 (18.5) |
| Fibrocavitary pattern | 60 (4.6) |
| Single nodule | 58 (4.4) |
| Hypersensitivity pneumonitis | 2 (0.2) |
| Others | 54 (4.1) |
| Observation period (month), mean (SD) | 60.9 ± 44.6 |

AIDS, acquired immunodeficiency syndrome; COI, cutoff index; COPD,
chronic obstructive pulmonary disease; ESR, erythrocyte sedimentation rate; NB, nodular bronchiectasis; SD, standard deviation

The median time from NTM-PD diagnosis to CPA diagnosis was 26 months (IQR, 12–58 months) (Figure 2). Of the 45 patients with NTM-PD infected with CPA, 35 were treated for CPA during the observation period (9 before and 26 after diagnosis of NTM-PD), and 5 patients required a change in NTM-PD medication for CPA treatment. The initial antifungal agents were voriconazole in 13 patients (28.9%) and itraconazole as well as micafungin in 6 patients (13.3%) (Table S2).

**FIGURE 2.**
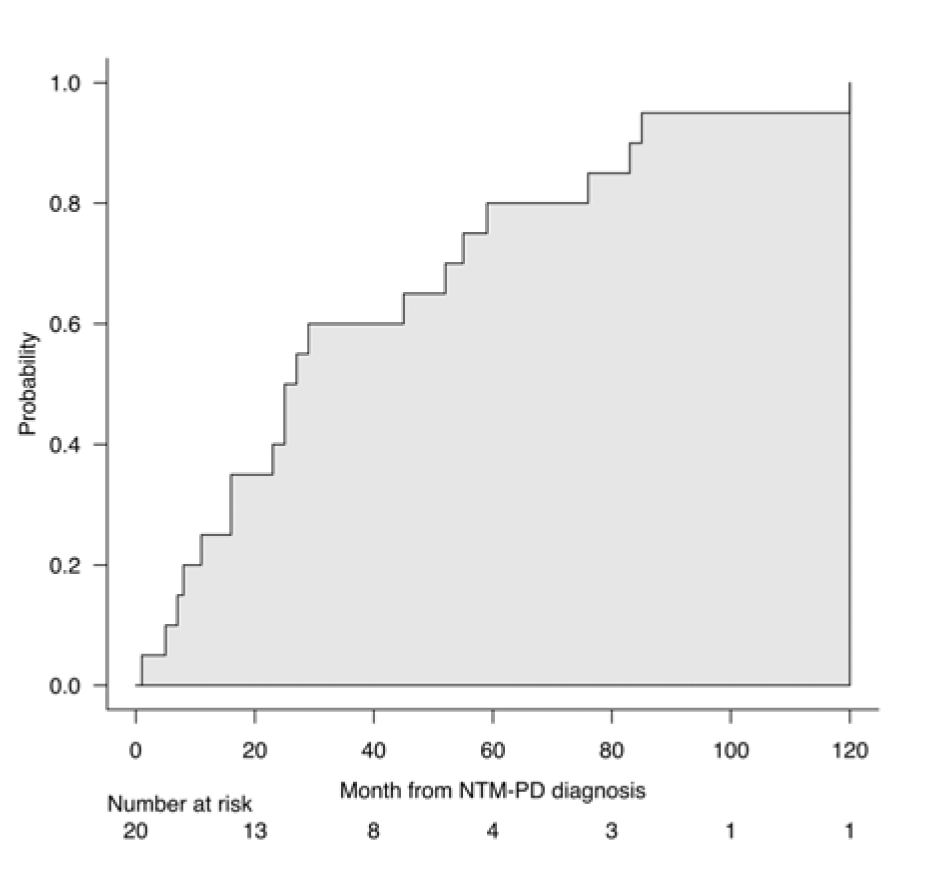
Duration from nontuberculous mycobacterial pulmonary disease diagnosis to chronic pulmonary aspergillosis diagnosis. Median duration from NTM-PD diagnosis to CPA diagnosis was 26 months (IQR, 12–58 months), with a mean of 38.4 ± 32.6 months. IQR, interquartile range; CPA, chronic pulmonary aspergillosis; NTM-PD, nontuberculous mycobacterial pulmonary disease

### Isolated Mycobacterial and *Aspergillus* Species

In this study, 1,328 strains were isolated and identified from 1,304 patients (Table S3). *Mycobacterium intracellulare* was the most common cause of NTM-PD (50.4%), followed by *M. avium* (40.5%), *Mycobacterium abscessus* (2.4%), *Mycobacterium kansasii* (2.4%), and *Mycobacterium avium-intracellulare* complex (1.7%). Of the 45 patients diagnosed with CPA, *Aspergillus* species were identified in 23 cases: *Aspergillus fumigatus* in 14 (60.1%), *Aspergillus niger* in 4 (17.4%), *Aspergillus terreus* in 3 (13.0%), and *Aspergillus flavus* and *Aspergillus nidulans* in one patient each (4.3%).

### Risk Factors for *Aspergillus* Coinfection

To examine the risk factors for NTM-PD with CPA, we compared the patient background at the time of new diagnosis of NTM-PD in 1,259 patients with NTM-PD without CPA and 45 patients with NTM-PD with CPA, out of 1,304 patients with confirmed NTM-PD. Univariate analysis revealed that male sex, low body mass index, smoking history, old tuberculosis, bronchiectasis, interstitial lung disease, COPD, pulmonary emphysema, chronic bronchitis, pulmonary aspergillosis, any dose of oral corticosteroids and immunosuppressants, low albuminemia, elevation of erythrocyte sedimentation rate, cavity formation, and radiological cavitary NB pattern and fibrocavitary pattern were risk factors for CPA complications. Multiple logistic regression analysis was carried out using age, male sex, COPD, oral corticosteroid use, and cavity formation. This analysis showed that male sex, COPD, oral corticosteroid use, and cavity formation were risk factors for CPA (Table 2, Table S4).

**TABLE 2.**
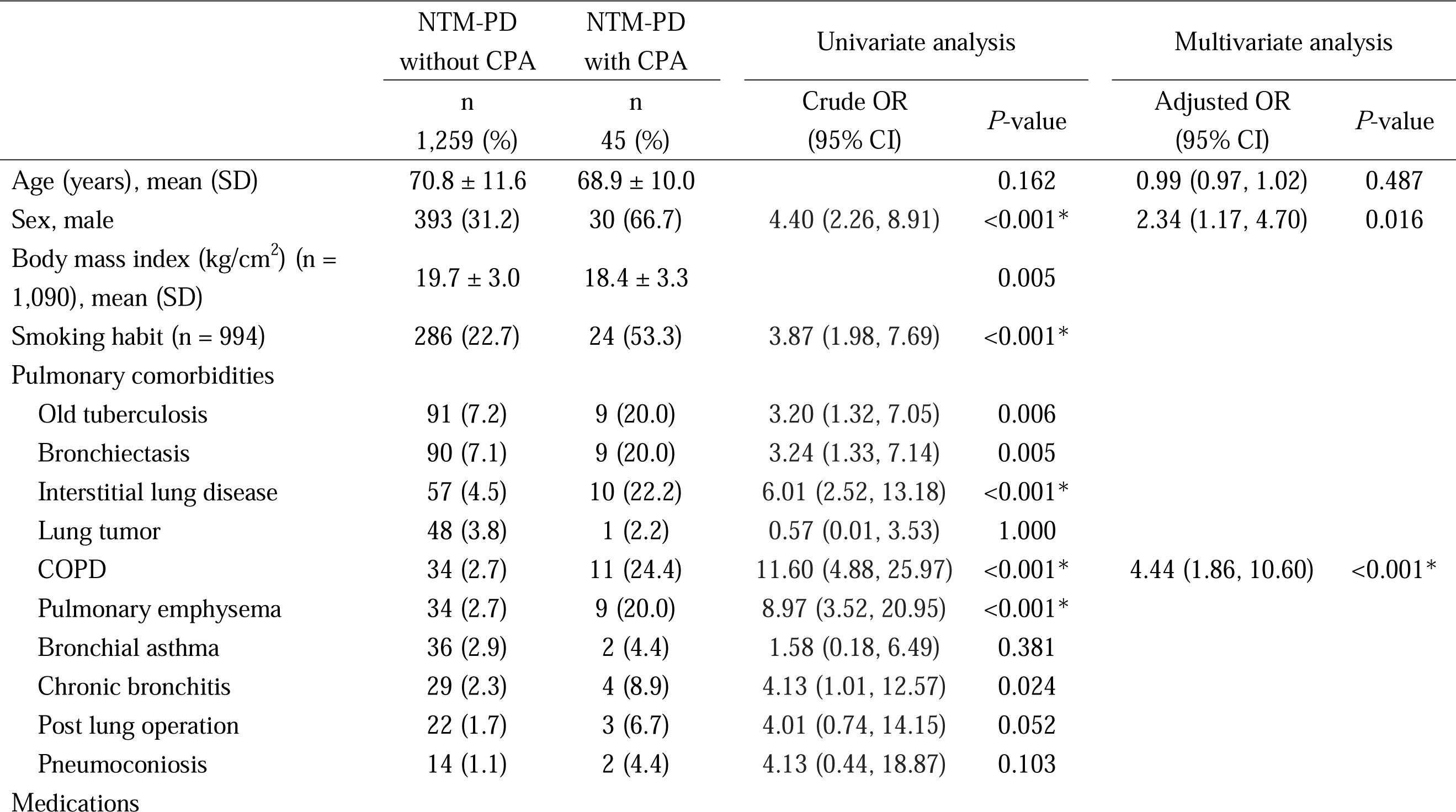
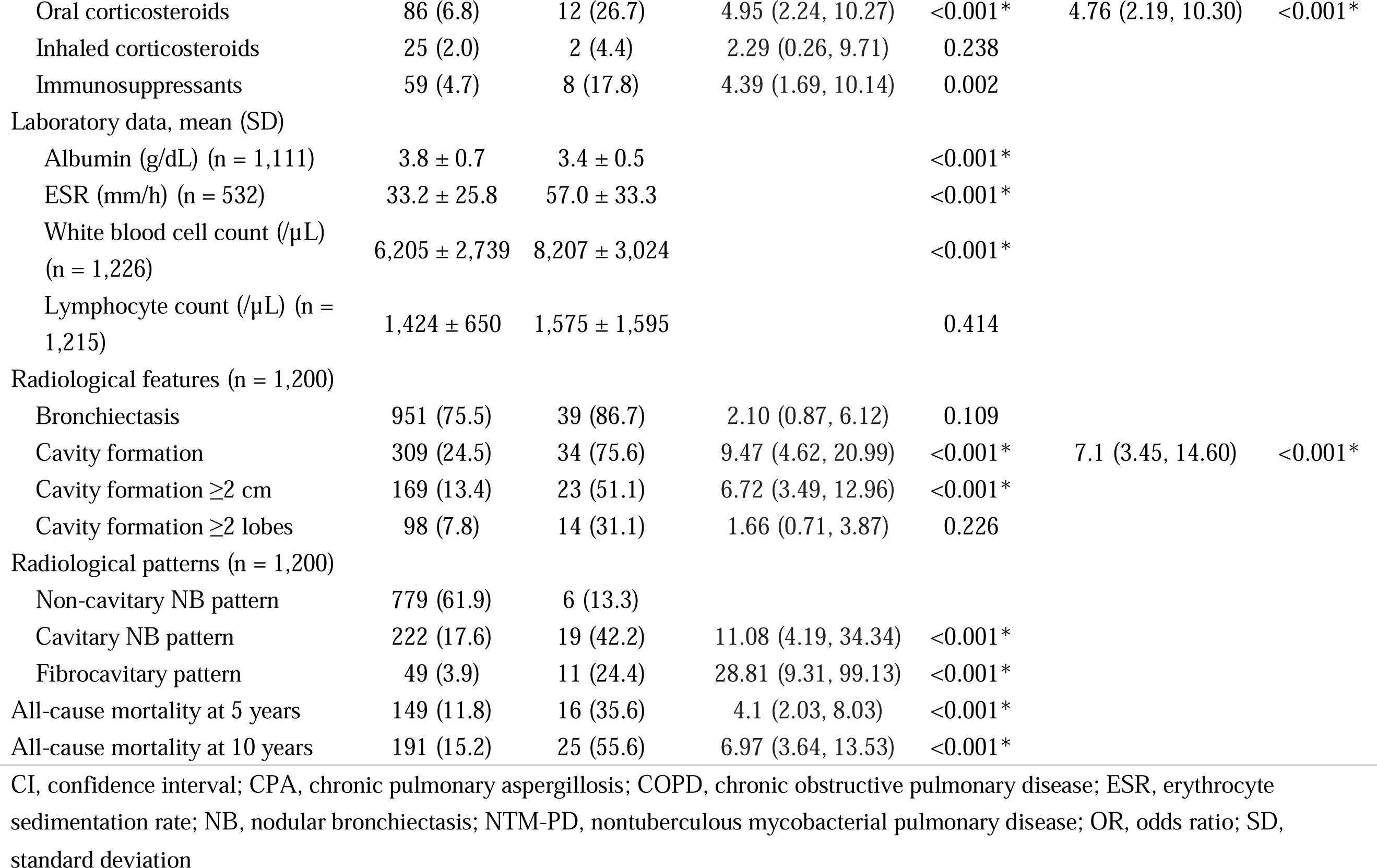
Characteristics of Patients With NTM-PD With or Without CPA.

|  | NTM-PD<br>without CPA | NTM-PD<br>with CPA | Univariate analysis |  | Multivariate analysis |  |
| --- | --- | --- | --- | --- | --- | --- |
|  | n<br>1,259 (%) | n<br>45 (%) | Crude OR<br>(95% CI) | *P*-value | Adjusted OR<br>(95% CI) | *P*-value |
| Age (years), mean (SD) | 70.8 ± 11.6 | 68.9 ± 10.0 |  | 0.162 | 0.99 (0.97, 1.02) | 0.487 |
| Sex, male | 393 (31.2) | 30 (66.7) | 4.40 (2.26, 8.91) | <0.001* | 2.34 (1.17, 4.70) | 0.016 |
| Body mass index (kg/cm<sup>2</sup>) (n = 1,090), mean (SD) | 19.7 ± 3.0 | 18.4 ± 3.3 |  | 0.005 |  |  |
| Smoking habit (n = 994) | 286 (22.7) | 24 (53.3) | 3.87 (1.98, 7.69) | <0.001* |  |  |
| Pulmonary comorbidities |  |  |  |  |  |  |
| Old tuberculosis | 91 (7.2) | 9 (20.0) | 3.20 (1.32, 7.05) | 0.006 |  |  |
| Bronchiectasis | 90 (7.1) | 9 (20.0) | 3.24 (1.33, 7.14) | 0.005 |  |  |
| Interstitial lung disease | 57 (4.5) | 10 (22.2) | 6.01 (2.52, 13.18) | <0.001* |  |  |
| Lung tumor | 48 (3.8) | 1 (2.2) | 0.57 (0.01, 3.53) | 1.000 |  |  |
| COPD | 34 (2.7) | 11 (24.4) | 11.60 (4.88, 25.97) | <0.001* | 4.44 (1.86, 10.60) | <0.001* |
| Pulmonary emphysema | 34 (2.7) | 9 (20.0) | 8.97 (3.52, 20.95) | <0.001* |  |  |
| Bronchial asthma | 36 (2.9) | 2 (4.4) | 1.58 (0.18, 6.49) | 0.381 |  |  |
| Chronic bronchitis | 29 (2.3) | 4 (8.9) | 4.13 (1.01, 12.57) | 0.024 |  |  |
| Post lung operation | 22 (1.7) | 3 (6.7) | 4.01 (0.74, 14.15) | 0.052 |  |  |
| Pneumoconiosis | 14 (1.1) | 2 (4.4) | 4.13 (0.44, 18.87) | 0.103 |  |  |
| Medications |  |  |  |  |  |  |
| Oral corticosteroids | 86 (6.8) | 12 (26.7) | 4.95 (2.24, 10.27) | <0.001* | 4.76 (2.19, 10.30) | <0.001* |
| Inhaled corticosteroids | 25 (2.0) | 2 (4.4) | 2.29 (0.26, 9.71) | 0.238 |  |  |
| Immunosuppressants | 59 (4.7) | 8 (17.8) | 4.39 (1.69, 10.14) | 0.002 |  |  |
| Laboratory data, mean (SD) |  |  |  |  |  |  |
| Albumin (g/dL) (n = 1,111) | 3.8 ± 0.7 | 3.4 ± 0.5 |  | <0.001* |  |  |
| ESR (mm/h) (n = 532) | 33.2 ± 25.8 | 57.0 ± 33.3 |  | <0.001* |  |  |
| White blood cell count (/μL) (n = 1,226) | 6,205 ± 2,739 | 8,207 ± 3,024 |  | <0.001* |  |  |
| Lymphocyte count (/μL) (n = 1,215) | 1,424 ± 650 | 1,575 ± 1,595 |  | 0.414 |  |  |
| Radiological features (n = 1,200) |  |  |  |  |  |  |
| Bronchiectasis | 951 (75.5) | 39 (86.7) | 2.10 (0.87, 6.12) | 0.109 |  |  |
| Cavity formation | 309 (24.5) | 34 (75.6) | 9.47 (4.62, 20.99) | <0.001* | 7.1 (3.45, 14.60) | <0.001* |
| Cavity formation ≥2 cm | 169 (13.4) | 23 (51.1) | 6.72 (3.49, 12.96) | <0.001* |  |  |
| Cavity formation ≥2 lobes | 98 (7.8) | 14 (31.1) | 1.66 (0.71, 3.87) | 0.226 |  |  |
| Radiological patterns (n = 1,200) |  |  |  |  |  |  |
| Non-cavitary NB pattern | 779 (61.9) | 6 (13.3) |  |  |  |  |
| Cavitary NB pattern | 222 (17.6) | 19 (42.2) | 11.08 (4.19, 34.34) | <0.001* |  |  |
| Fibrocavitary pattern | 49 (3.9) | 11 (24.4) | 28.81 (9.31, 99.13) | <0.001* |  |  |
| All-cause mortality at 5 years | 149 (11.8) | 16 (35.6) | 4.1 (2.03, 8.03) | <0.001* |  |  |
| All-cause mortality at 10 years | 191 (15.2) | 25 (55.6) | 6.97 (3.64, 13.53) | <0.001* |  |  |

CI, confidence interval; CPA, chronic pulmonary aspergillosis; COPD, chronic obstructive pulmonary disease; ESR, erythrocyte sedimentation rate; NB, nodular bronchiectasis; NTM-PD, nontuberculous mycobacterial pulmonary disease; OR, odds ratio; SD,
standard deviation

### Survival Analysis for All-Cause Mortality

Among patients with NTM-PD without CPA (n = 1,259) and with CPA (n = 45), all-cause mortality from diagnosis of NTM-PD to the date of last observation was 15.3% (n = 192) and 55.6% (n = 25), respectively (log-rank test *P* < .001; crude HR, 3.98; 95% CI, 2.62–6.05). Patients with CPA had significantly worse prognoses than those without CPA (Table 3). Kaplan–Meier curves are shown in Figure 3A. To determine whether the presence of CPA was an independent poor prognostic factor or whether the patient’s predisposition to CPA was a poor prognostic factor, we performed propensity score matching for age, male sex, COPD, oral corticosteroid use, and cavity formation. The five values used for matching had a standardized mean difference of <0.10 (Table S5). The results showed that CPA was a significant poor prognostic factor (Figure 3B) (log-rank test *P* = .036; adjusted HR, 1.59; 95% CI, 0.84–3.02) (Table 3).

**FIGURE 3.**
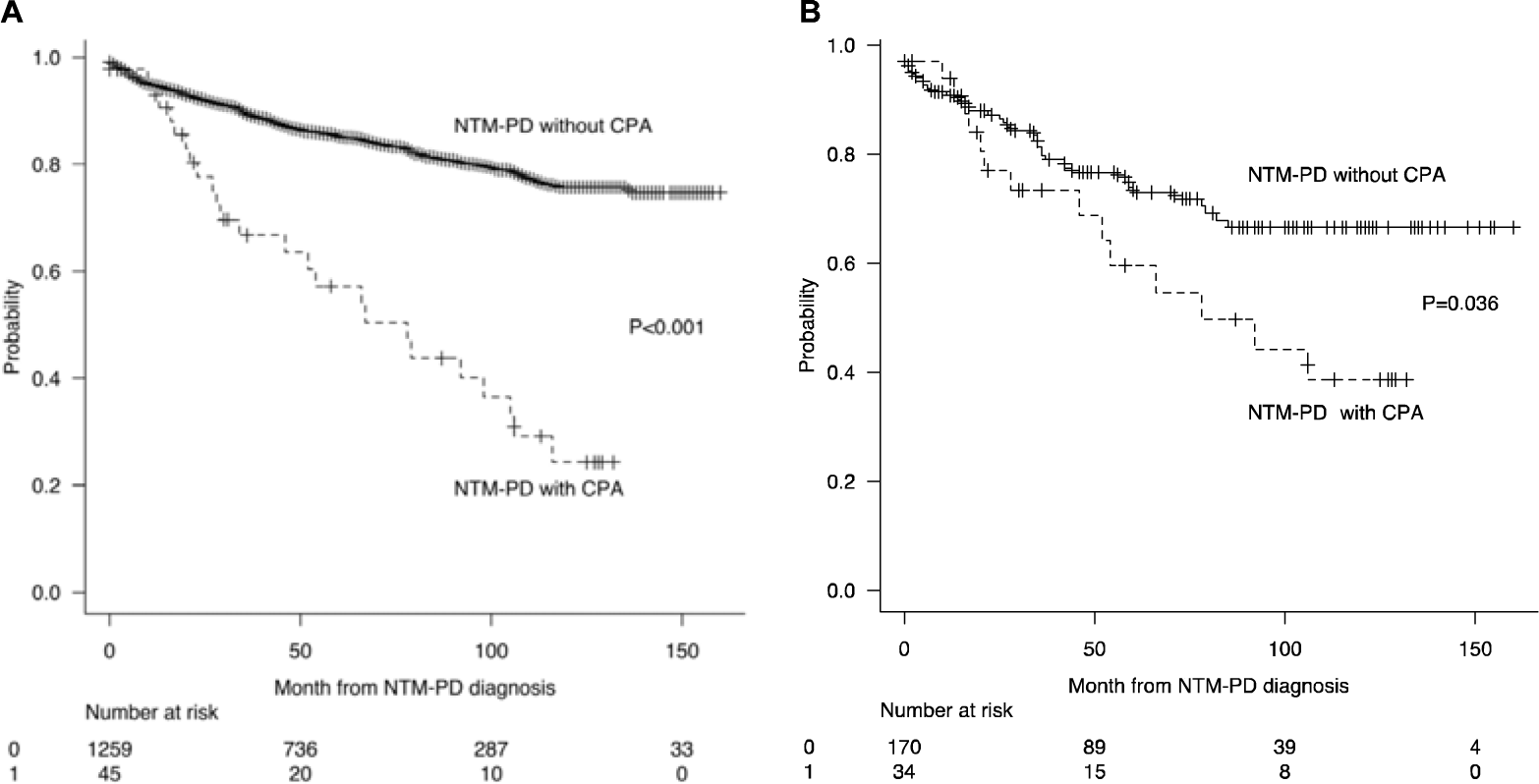
Kaplan–Meier analysis for patients with NTM-PD with or without CPA, before (A) and after (B) propensity score matching. Patients with NTM-PD infected with CPA had significantly worse prognoses than those without CPA (A). Analysis after propensity score matching for age, male sex, COPD, oral corticosteroid use, and cavity formation showed that CPA was a significant poor prognostic factor (B). COPD, chronic obstructive pulmonary disease; CPA, chronic pulmonary aspergillosis; NTM-PD, nontuberculous mycobacterial pulmonary disease

**TABLE 3.** Comparison of All-Cause Mortality.

|  | Before propensity score matching |  | After propensity score matching |  |
| --- | --- | --- | --- | --- |
|  | Number of events/All cases | Crude HR (95% CI) | Number of events/All cases | Adjusted HR (95% CI) |
| NTM-PD without CPA | 192/1,259 | reference | 43/170 | reference |
| NTM-PD with CPA | 25/45 | 3.98<br>(2.62–6.05) | 15/34 | 1.59<br>(0.84–3.02) |

CI, confidence interval; CPA, chronic pulmonary aspergillosis; HR, hazards ratio; NTM-PD, nontuberculous mycobacterial pulmonary disease

## DISCUSSION

The results of this multicenter study provide significant insights into CPA coinfection in patients with NTM-PD. Our key findings include the CPA coinfection rate, identification of specific risk factors, and demonstration of its impact on long-term prognosis.

The observed CPA coinfection rate in our cohort was lower than that reported in previous observational studies (3.5% vs. 3.9–11.0%).^7-9,11,17,18^ This discrepancy may be attributed to the heterogeneity of medical facilities and variability in CPA diagnostic practices. The aforementioned studies were conducted at single-center hospitals, potentially resulting in a substantial number of patients with NTM-PD with complex disease states, owing to selection bias. Our study included 1,304 patients from 18 medical facilities, including non-tertiary care centers, potentially encompassing representative patients with NTM-PD, including mild cases. Furthermore, the diagnosis of CPA is sometimes challenging because the detection rate of fungal microscopy and fungal culture of respiratory specimens is low, β-1,3-D-glucan is not specific for *Aspergillus* in serological tests, galactomannan antigen is not sensitive, and anti-*Aspergillus* IgG antibodies are specific but not sensitive, particularly for non-fumigatus species.^19^ This implies that the diagnostic accuracy of CPA varies among facilities. However, the impact is likely to be small because all facilities participating in this study were able to test for bronchoscopy, β-1,3-D-glucan, galactomannan antigen, and antibodies. We have also previously reported on NTM-PD and CPA complications based on Japanese large-scale claims database.^20^ The reported incidence of CPA for NTM-PD was 2.29%, which is lower than that reported in the present study. However, the previous report included patients treated for both NTM-PD and CPA, whereas the coinfection rate in the current report included all patients, regardless of treatment, and thus better reflects the real-world coinfection rate. In this study, *M. intracellulare* and *M. avium* were the isolated dominant species. *Aspergillus fumigatus* was the most common species causing CPA, followed by *A. niger*. This microbiological epidemiology is similar to that of previous reports^21^; however, *Aspergillus* colonization was excluded from our study. The treatment of patients with NTM-PD coinfected with CPA is challenging, owing to drug-drug interactions.^10,21,22^ In our study, in five patients with NTM-PD coinfected with CPA, treatment had to be changed during the observation period. Physicians should carefully consider the risk of CPA coinfection in patients with NTM-PD.

Previous studies have reported the following risk factors for CPA coinfection in patients with NTM-PD: fungal balls and cavities with adjacent extrapleural fat, systemic steroids, cavity formation, emphysema, hypoalbuminemia, older age, male sex, COPD, and *M. abscessus* complex.^7–9,11,17^ In our study, we demonstrated that male sex, COPD, oral corticosteroid use, and cavity formation are risk factors for CPA complications. These factors may help clinicians identify patients with NTM-PD at a high risk of developing CPA and thereby lead to early CPA diagnosis.

Previous reports have not adequately evaluated patient background to determine whether CPA complication is an independent poor prognostic factor or whether mortality is high in populations at a high risk of CPA complications owing to case number limitations. We confirmed that CPA comorbidity was associated with a poor long-term prognosis in patients with NTM-PD, even after adjusting for confounding factors through propensity score matching. This result corroborates the findings from our previous analysis using the Japanese claims database, which examined cases requiring treatment for both NTM-PD and CPA coinfection.^20^ This highlights the importance of vigilant monitoring of CPA in patients with NTM-PD, particularly in those with identified risk factors.

The strengths of this study include its large sample size, multicenter design, and long-term follow-up. Our study provides valuable insights; however, several limitations should be acknowledged. First, this study was retrospective, and unlike prospective studies, selection bias, data quality and completeness, and confounding variables should be considered as potential limitations. Second, there is potential variability in the practice of CPA diagnosis. CPA diagnosis is sometimes challenging, and the design of a multicenter study using medical records can exhibit considerable variation. Third, generalizability outside Japan is limited because the etiology of NTM differs between Japan and other areas.^23,24^ However, *M. avium* complex was the predominant organism in this study. In many other areas, *M. avium* complex is one of the primary NTM-PD causative species, albeit with some regional variation. Fourth, our study included 45 cases of CPA. However, no CPA subtype analysis was performed. Systematic reviews and meta-analyses of CPA have demonstrated that CPA subtype is associated with mortality and that patients with CCPA, CFPA, and SAIA show worse prognoses than those with simple aspergilloma.^25^ Nevertheless, in our study, 42 patients were diagnosed with CCPA, CFPA, or SAIA. Consequently, the impact on outcome was considered minimal.

In conclusion, the findings of this study suggest that male sex, COPD, oral corticosteroid use, and cavity formation are risk factors for CPA coinfection in patients with NTM-PD. Additionally, CPA is an independent poor prognostic factor. Clinicians must consider CPA when treating patients with NTM-PD, particularly those with risk factors. Extended observational studies of prospective trials are necessary to elucidate the risk factors and long-term prognosis of patients with NTM-PD and CPA coinfection.

## Abbreviations

CCPA: chronic cavitary pulmonary aspergillosis
CFPA: chronic fibrosing pulmonary aspergillosis
CPPA: chronic progressive pulmonary aspergillosis
CPA: chronic pulmonary aspergillosis
JSTNM: Japanese Society for Tuberculosis and Nontuberculous Mycobacteriosis
NB: nodular bronchiectasis
NTM-PD: nontuberculous mycobacterial pulmonary disease
SAIA: subacute invasive pulmonary aspergillosis

## ACKNOWLEDGMENTS

The authors are grateful to Y. Ito, S. Koga, H. Ashizawa, K. Fukushima, N. Matsuo, S. Yoshioka, D. Noritomi, Y. Fukushima, S. Kaneko, R. Morio, R. Mizuta, T. Inoue, T. Ikeda, A. Hara, D. Setoguchi, K. Mine, Y. Hirano, Y. Nagayoshi, R. Morishita, Y. Usui, K. Yoshiyama, S. Tomari, S. Doi, A. Umemura, Y. Umeyama, T. Miyamura, Ryosuke Ogata, Ryo Ogata, C. Iketani, K. Nemoto, M. Funada, Y. Isoshima, S. Shigemi, H. Kanda, M. Sumiyoshi, E. Kitamura, A. Kitamura, N. Matsumoto, A. Sano, A. Matsuo, E. Mitsutome, Y. Ideguchi, M. Yamasue, R. Takaki, and K. Tobino for their review of the medical records. We also thank R. Kawasaki and H. Yano of Clinical Research Center, Nagasaki University Hospital for building an electronic data-capture system; S. Morimoto of Clinical Research Center, Nagasaki University Hospital, for advice on formal analysis; and Editage (www.editage.jp) for English language editing. This work was partly conducted by Non-profit Organization Aimed to Support Community Medicine Research in Nagasaki (3612) and the joint research program of the Research Center for GLOBAL and LOCAL Infectious Diseases, Oita University (2023B15). The abstract of this manuscript was presented at the 99th Congress of the Japanese Society of Tuberculosis and Non-Tuberculous Mycobacteriosis on May 31, 2024, and the 64th annual meeting of the Japanese Respiratory Society on April 5, 2024.

## AUTHOR CONTRIBUTIONS

**Yasuhiro Tanaka:** Investigation, formal analysis, discussion of results, and writing - original draft preparation. **Shotaro Ide:** Conceptualization, data curation, formal analysis, funding acquisition, investigation, methodology, project administration, visualization, writing - original draft preparation, and writing - review & editing. **Takahiro Takazono:** Conceptualization, methodology, project administration, and writing - review & editing. **Kazuaki Takeda:** Formal analysis, investigation, methodology, resources, and writing - review & editing. **Naoki Iwanaga:** Investigation and writing - review & editing. **Masataka Yoshida:** Investigation, and writing - review & editing. **Naoki Hosogaya:** Data curation, resources, and writing - review & editing. **Yusei Tsukamoto:** Investigation and writing - review & editing. **Satoshi Irifune:** Investigation and writing - review & editing. **Takayuki Suyama:** Investigation and writing - review & editing. **Tomo Mihara:** Investigation and writing - review & editing. **Akira Kondo:** Investigation and writing - review & editing. **Tsutomu Kobayashi:** Investigation and writing - review & editing. **Yuichi Fukuda:** Investigation and writing - review & editing. **Eisuke Sasaki:** Investigation and writing - review & editing. **Toyomitsu Sawai:** Investigation and writing - review & editing. **Yasuhito Higashiyama:** Investigation and writing - review & editing. **Kohji Hashiguchi:** Investigation and writing - review & editing. **Minako Hanaka:** Investigation and writing - review & editing. **Toshihiko Ii:** Investigation and writing - review & editing. **Kiyoyasu Fukushima:** Investigation and writing - review & editing. **Kosaku Komiya:** Funding acquisition, investigation, and writing - review & editing. **Taiga Miyazaki:** Investigation and writing - review & editing. **Katzuhiro Yatera:** Investigation and writing - review & editing. **Koichi Izumikawa:** Methodology and writing - review & editing. **Akitsugu Furumoto:** Methodology and writing - review & editing. **Katsunori Yanagihara:** Resources and writing - review & editing. Hiroshi Mukae: Conceptualization, supervision, and writing - review & editing.

## DATA AVAILABILITY STATEMENT

The datasets analyzed in this study are not publicly available.

## HUMAN AND ETHICS APPROVAL DECLARATION

This study was conducted in accordance with the guidelines of the Declaration of Helsinki, and approved by the appropriate Institutional Review Board of Nagasaki University Hospital (approval number: 22121903). Verbal informed consent was obtained whenever possible, and as this was a retrospective observational study, an opt-out procedure was provided to patients.

