## Supplementary material for "Risk Factors and Long-term Prognosis for Coinfection of Nontuberculous Mycobacterial Pulmonary Disease and Chronic Pulmonary Aspergillosis: A Multicenter Observational Study in Japan": Table S1

| **Table S1. List of Hospitals Participating in the Study** | | | | | |
| --- | --- | --- | --- | --- | --- |
|  | Hospital | Prefecture | Number of beds in 2022 | NTM-PD without CPA n (%) | NTM-PD with CPA n (%) |
| 1 | Nagasaki University Hospital | Nagasaki | 874 | 116 (88.5) | 15 (11.5) |
| 2 | Nagasaki Harbor Medical Center | Nagasaki | 513 | 48 (100.0) | 0 (0.0) |
| 3 | Japanese Red Cross Nagasaki Genbaku Hospital | Nagasaki | 315 | 40 (93.0) | 3 (7.0) |
| 4 | NHO Nagasaki Medical Center | Nagasaki | 643 | 98 (97.0) | 3 (3.0) |
| 5 | Japanese Red Cross Nagasaki Genbaku Isahaya Hospital | Nagasaki | 130 | 96 (100.0) | 0 (0.0) |
| 6 | Japan Community Health care Organization Isahaya General Hospital | Nagasaki | 325 | 128 (96.2) | 5 (3.8) |
| 7 | Nagasaki Prefecture Shimabara Hospital | Nagasaki | 254 | 113 (98.3) | 2 (1.7) |
| 8 | Izumikawa Hospital | Nagasaki | 120 | 36 (100.0) | 0 (0.0) |
| 9 | Sasebo City General Hospital | Nagasaki | 594 | 151 (95.0) | 8 (5.0) |
| 10 | Sasebo Chuo Hospital | Nagasaki | 312 | 57 (98.3) | 1 (1.7) |
| 11 | Hokusho Central Hospital | Nagasaki | 189 | 16 (100.0) | 0 (0.0) |
| 12 | Nagasaki Goto Chuoh Hospital | Nagasaki | 304 | 46 (97.9) | 1 (2.1) |
| 13 | Department of Respiratory Medicine, University of Occupational and Environmental Health, Japan | Fukuoka | 678 | 22 (100.0) | 0 (0.0) |
| 14 | University of Miyazaki Hospital | Miyazaki | 632 | 40 (90.9) | 4 (9.1) |
| 15 | NHO Miyazaki Higashi Hospital | Miyazaki | 250 | 71 (98.6) | 1 (1.4) |
| 16 | NHO Ureshino Medical Center | Saga | 399 | 69 (98.6) | 1 (1.4) |
| 17 | Oita University Hospital | Oita | 604 | 43 (100.0) | 0 (0.0) |
| 18 | Iizuka Hospital | Fukuoka | 1,048 | 69 (98.6) | 1 (1.4) |
|  | Overall |  | 8,184 | 1,259 (96.5) | 45 (3.5) |
| NTM-PD, nontuberculous mycobacterial pulmonary disease; CPA, chronic pulmonary aspergillosis | | | | | |
