## Supplementary material for "Risk Factors and Long-term Prognosis for Coinfection of Nontuberculous Mycobacterial Pulmonary Disease and Chronic Pulmonary Aspergillosis: A Multicenter Observational Study in Japan": Table S2

| **Table S2. Treatment for Chronic Pulmonary Aspergillosis** | |
| --- | --- |
|  | n = 45 (%) |
| CPA treatment before NTM-PD diagnosis | 9 (20.0) |
| CPA treatment after NTM-PD diagnosis | 26 (57.8) |
| No treatment for CPA | 10 (22.2) |
| Antifungal agents at initial therapy |  |
| Voriconazole | 13 (28.9) |
| Micafungin | 6 (13.3) |
| Itraconazole | 6 (13.3) |
| Caspofungin | 2 (4.4) |
| Liposomal amphotericin B | 1 (2.2) |
| Amphotericin B | 1 (2.2) |
| Unknown | 6 (13.3) |
| CPA, chronic pulmonary aspergillosis; NTM-PD, nontuberculous mycobacterial pulmonary disease | |
