## Supplementary material for "Risk Factors and Long-term Prognosis for Coinfection of Nontuberculous Mycobacterial Pulmonary Disease and Chronic Pulmonary Aspergillosis: A Multicenter Observational Study in Japan": Table S3

| **Table S3. Isolated Mycobacterial species** | |
| --- | --- |
|  | n (%) |
| All isolated strains | 1,328 (100) |
| *Mycobacterium intracellulare* | 669 (50.4) |
| *Mycobacterium avium* | 538 (40.5) |
| *Mycobacterium avium-intracellulare* complex | 23 (1.7) |
| *Mycobacterium abscessus* | 32 (2.4) |
| *Mycobacterium kansasii* | 32 (2.4) |
| *Mycobacterium gordonae* | 11 (0.8) |
| *Mycobacterium chelonae* | 5 (0.4) |
| *Mycobacterium fortuitum* | 2 (0.2) |
| *Mycobacterium scrofulaceum* | 2 (0.2) |
| *Mycobacterium paragornae* | 1 (0.1) |
| *Mycobacterium nonchromogenicum* | 1 (0.1) |
| *Mycobacterium xenopi* | 1 (0.1) |
| *Mycobacterium szulgai* | 1 (0.1) |
| *Mycobacterium marinum* | 1 (0.1) |
| Not identified | 9 (0.7) |
