## Supplementary material for "Risk Factors and Long-term Prognosis for Coinfection of Nontuberculous Mycobacterial Pulmonary Disease and Chronic Pulmonary Aspergillosis: A Multicenter Observational Study in Japan": Table S4

| **Table S4. All Characteristics of Patients With Nontuberculous Mycobacterial Pulmonary Disease With or Without Chronic Pulmonary Aspergillosis** | | | | | |
| --- | --- | --- | --- | --- | --- |
|  | NTM-PD  without CPA | NTM-PD with CPA |  | Univariative analysis | |
|  | n | n |  | Crude OR (95% CI) | *P*-value |
|  | 1 259 (%) | 45 (%) |  |  |  |
| Age (years), mean (SD) | 70.8 ± 11.6 | 68.9 ± 10.0 |  |  | 0.162 |
| Sex, male | 393 (31.2) | 30 (66.7) |  | 4.40 (2.26, 8.91) | <0.001 |
| Body mass index (n = 1,090), mean (SD) | 19.7 ±3.0 | 18.4 ± 3.3 |  |  | 0.005 |
| Smoking habit (n = 994) | 286 (22.7) | 24 (53.3) |  | 3.87 (1.98, 7.69) | <0.001 |
| Pulmonary comorbidities |  |  |  |  |  |
| Old tuberculosis | 91 (7.2) | 9 (20.0) |  | 3.20 (1.32, 7.05) | 0.006 |
| Bronchiectasis | 90 (7.1) | 9 (20.0) |  | 3.24 (1.33, 7.14) | 0.005 |
| Interstitial lung disease | 57 (4.5) | 10 (22.2) |  | 6.01 (2.52, 13.18) | <0.001 |
| Lung tumor | 48 (3.8) | 1 (2.2) |  | 0.57 (0.01, 3.53) | 1.000 |
| COPD | 34 (2.7) | 11 (24.4) |  | 11.60 (4.88, 25.97) | <0.001 |
| Pulmonary emphysema | 34 (2.7) | 9 (20.0) |  | 8.97 (3.52, 20.95) | <0.001 |
| Bronchial asthma | 36 (2.9) | 2 (4.4) |  | 1.58 (0.18, 6.49) | 0.381 |
| Chronic bronchitis | 29 (2.3) | 4 (8.9) |  | 4.13 (1.01, 12.57) | 0.024 |
| Post lung operation | 22 (1.7) | 3 (6.7) |  | 4.01 (0.74, 14.15) | 0.052 |
| Pneumoconiosis | 14 (1.1) | 2 (4.4) |  | 4.13 (0.44, 18.87) | 0.103 |
| Pulmonary aspergillosis | 2 (0.2) | 11 (24.4) |  | 199.38 (41.10, 1930.14) | <0.001 |
| Chronic respiratory failure with home oxygen therapy | 3 (0.2) | 1 (2.2) |  | 9.59 (0.179, 122.32) | 0.130 |
| Asbestos-related pleural disease | 3 (0.2) | 1 (2.2) |  | 9.47 (0.18, 120.51) | 0.131 |
| Pulmonary sarcoidosis | 1 (0.1) | 0 (0.0) |  | 0.00 (0.00, 1072.86) | 1.000 |
| Systemic comorbidities |  |  |  |  |  |
| Solid tumor, localized | 156 (12.4) | 4 (8.9) |  | 0.69 (0.18, 1.94) | 0.645 |
| Connective tissue diseases | 105 (8.3) | 8 (17.8) |  | 2.37 (0.93, 5.36) | 0.051 |
| Diabete mellitus | 93 (7.4) | 4 (8.9) |  | 1.22 (0.31, 3.49) | 0.572 |
| Cerebrovascular disease | 44 (3.5) | 3 (6.7) |  | 1.97 (0.38, 6.56) | 0.219 |
| Liver disease, moderate/severe | 42 (3.3) | 1 (2.2) |  | 0.66 (0.016, 4.07) | 1.000 |
| Congestive heart failure | 31 (2.5) | 2 (4.4) |  | 1.84 (0.21, 7.65) | 0.316 |
| Solid tumor, metastatic | 32 (2.5) | 1 (2.2) |  | 0.87 (0.02, 5.48) | 1.000 |
| Myocardial infarction | 30 (2.4) | 1 (2.2) |  | 0.93 (0.02, 5.88) | 1.000 |
| Dementia | 29 (2.3) | 0 (0.0) |  | 0.00 (0.00, 3.86) | 0.621 |
| Lymphoma | 23 (1.8) | 1 (2.2) |  | 1.22 (0.03, 7.88) | 0.573 |
| Peptic ulcer disease | 17 (1.4) | 2 (4.4) |  | 3.39 (0.37, 15.04) | 0.137 |
| Renal disease | 16 (1.3) | 2 (4.4) |  | 3.61 (0.39, 16.13) | 0.126 |
| Peripheral vascular disease | 7 (0.6) | 0 (0.0) |  | 0.00 (0.00, 19.88) | 1.000 |
| Leukemia | 5 (0.4) | 0 (0.0) |  | 0.00 (0.00, 31.18) | 1.000 |
| AIDS | 3 (0.2) | 0 (0.0) |  | 0.00 (0.00, 68.72) | 1.000 |
| Paralysis | 2 (0.2) | 0 (0.0) |  | 0.00 (0.00, 150.37) | 1.000 |
| Medications for comorbidities |  |  |  |  |  |
| Oral corticosteroids | 86 (6.8) | 12 (26.7) |  | 4.95 (2.24, 10.27) | <0.001 |
| Inhaled corticosteroids | 25 (2.0) | 2 (4.4) |  | 2.29 (0.26, 9.71) | 0.238 |
| Immunosuppressants | 59 (4.7) | 8 (17.8) |  | 4.39 (1.69, 10.14) | 0.002 |
| Chemotherapy | 17 (1.4) | 1 (2.2) |  | 1.66 (0.04, 11.07) | 0.471 |
| Biological agents | 7 (0.6) | 1 (2.2) |  | 4.06 (0.09, 32.71) | 0.246 |
| Laboratory Data, mean (SD) |  |  |  |  |  |
| Albumin (g/dL) (n = 1,111) | 3.8 ± 0.7 | 3.4 ± 0.5 |  |  | <0.001 |
| ESR (mm/h) (n = 532) | 33.2 ± 25.8 | 57.0 ± 33.3 |  |  | <0.001 |
| White blood cell count (/µL) (n = 1,226) | 6,205 ± 2,739 | 8,207 ± 3,024 |  |  | <0.001 |
| Lymphocyte count (/µL) (n = 1,215) | 1,424 ± 650 | 1,575 ± 1,595 |  |  | 0.414 |
| Galactomannan antigen (COI) (n = 303) | 0.71 ± 0.91 | 0.88 ± 1.13 |  |  | 0.834 |
| Radiological Features (n = 1,200) |  |  |  |  |  |
| Bronchiectasis | 951 (75.5) | 39 (86.7) |  | 2.10 (0.87, 6.12) | 0.109 |
| Cavity formation | 309 (24.5) | 34 (75.6) |  | 9.47 (4.62, 20.99) | <0.001 |
| Cavity formation ≥ 2 cm | 169 (13.4) | 23 (51.1) |  | 6.72 (3.49, 12.96) | <0.001 |
| Cavity formation ≥ 2 lobes | 98 (7.8) | 14 (31.1) |  | 1.66 (0.71, 3.87) | 0.226 |
| Radiological Patterns (n = 1,200) |  |  |  |  |  |
| Non-cavitary NB pattern | 779 (61.9) | 6 (13.3) |  | reference |  |
| Cavitary NB pattern | 222 (17.6) | 19 (42.2) |  | 11.08 (4.19, 34.34) | <0.001 |
| Fibrocavitary pattern | 49 (3.9) | 11 (24.4) |  | 28.81 (9.31, 99.13) | <0.001 |
| Single nodule | 57 (4.5) | 1 (2.2) |  | 2.27 (0.05, 19.25) | 0.394 |
| Hypersensitivity pneumonitis | 2 (0.2) | 0 (0.0) |  | 0.00 (0.00, 753.75) | 1.000 |
| Others | 50 (4.0) | 4 (8.9) |  | 10.32 (2.07, 45.13) | 0.002 |
| Observation period (month), mean (SD) | 61.2 ± 44.6 | 53.8 ± 43.0 |  |  | 0.346 |
| All-cause mortality at 5 years | 149 (11.8) | 16 (35.6) |  | 4.1 (2.03, 8.03) | <0.001 |
| All-cause mortality at 10 years | 191 (15.2) | 25 (55.6) |  | 6.97 (3.64, 13.53) | <0.001 |
| COI, cutoff index; NB, nodular bronchiectatic | | | | | |
