## Supplementary material for "Risk Factors and Long-term Prognosis for Coinfection of Nontuberculous Mycobacterial Pulmonary Disease and Chronic Pulmonary Aspergillosis: A Multicenter Observational Study in Japan": Table S5

| **Table S5. Characteristics of Patients With Nontuberculous Mycobacterial Pulmonary Disease With or Without Chronic Pulmonary Aspergillosis Before And After Propensity Score Matching** | | | | | | | | | | | |
| --- | --- | --- | --- | --- | --- | --- | --- | --- | --- | --- | --- |
|  | Before propensity score matching | | | | |  | | After propensity score matching | | | |
|  | NTM-PD  without CPA | NTM-PD  with CPA | *P*-value | SMD |  | | NTM-PD  without CPA | | NTM-PD  with CPA | *P*-value | SMD |
|  | n = 1,259 (%) | n = 45 (%) |  |  |  | | n = 170 (%) | | n = 34 (%) |  |  |
| Age, mean (SD) | 70.8 ± 11.6 | 68.9±9.95 | <0.001 | 0.669 |  | | 71.0 ± 9.00 | | 71.0 ± 8.92 | 0.75 | 0.058 |
| Sex, male | 393 (31.2) | 30 (66.7) | <0.001 | 0.758 |  | | 100 (58.8) | | 20 (58.8) | 1.00 | <0.001 |
| COPD | 34 (2.7) | 11 (24.4) | <0.001 | 0.669 |  | | 12 (7.1) | | 3 (8.8) | 0.72 | 0.065 |
| Oral corticosteroids | 86 (6.8) | 12 (26.7) | <0.001 | 0.551 |  | | 24 (14.1) | | 4 (11.8) | 1.00 | 0.07 |
| Cavity formation | 309 (24.6) | 34 (75.6) | <0.001 | 1.186 |  | | 124 (72.9) | | 25 (73.5) | 1.00 | 0.013 |
| SMD, standardised mean difference | | | | | | | | | | | |
